# Small Enough to Use, Sharper Together: VQIDS and FIRM in U.S. Youth Ages 6–18

**DOI:** 10.64898/2026.10.13.26365411

**Authors:** Guillermo Perez Algorta, Lilian Y. Li, Yinuo Liu, Eric A. Youngstrom, Phoebe A. Rodda, A. John Rush

**Author notes:** Correspondence concerning this article should be addressed to Eric A. Youngstrom, Institute for Mental and Behavioral Health Research, Nationwide Children’s Hospital and Department of Psychiatry, 444 Butterfly Gardens Drive, Columbus, OH, 43215.

## Abstract

**Objective:** Brief measures of depressive symptoms and family psychiatry history may support the identification of youth who warrant further assessment, but their interpretation requires population-based measurement evidence. This study evaluated the Very Quick Inventory of Depressive Symptomatology (VQIDS) and Family Index of Risk for Mood (FIRM) in the BENCHMARK 2025 national survey.

**Method:** The sample consisted of 2,252 parent reports for children aged 6-18 years and 1,008 linked adolescent self-reports aged 12-18 years. Analyses examined VQIDS dimensionality, reliability, item functioning, age- and sex-based norms, and informant agreement, alongside FIRM score distributions, alternative scoring approaches, and associations with depressive symptoms.

**Results:** VQIDS findings supported a single dimension and good internal consistency, with greatest IRT precision at elevated symptom levels and no practically meaningful differential item functioning across age or sex. Two-week stability was moderate, and parent-adolescent agreement was lower within more symptomatic subsets. FIRM totals showed stronger cross-informant agreement and retest stability. Family history scores were associated with higher VQIDS percentiles and discriminated scores at or above the 90^th^ percentile across informants (AUROC = .670-.698). FIRM’s associations were strongest with internalizing symptoms, although evidence was mixed for mood specificity.

**Conclusions:** These findings provide a measurement and normative foundation for brief depressive symptom assessment and support family history as a complementary correlate of symptom elevation. Diagnostic and prospective prediction utility require further evaluation.

## VQIDS Measurement and Norms, and FIRM Characterization

Depression often emerges during adolescence and can disrupt functioning during this critical developmental period (Kessler et al., 2005; Thapar et al., 2012). In recent years, national surveys document rising rates of adolescent depressive symptoms and episodes, yet many affected youth remain unidentified or untreated (Mojtabai et al., 2016; SAMHSA, 2023). These gaps have motivated screening in settings that routinely serve young people, including primary care and school-based universal screening (Sekhar et al., 2021; Zuckerbrot et al., 2018). Brief, interpretable measures may help extend these efforts within the time and resource constraints of these services.

Family psychiatric history provides complementary information about mood vulnerability. Depression has a substantial heritable component, and children of parents with mood disorders are at elevated risk for mental health difficulties (Rasic et al., 2014; Sullivan et al., 2000). Accordingly, clinical guidelines recognize family history as relevant to identifying adolescents who may require closer monitoring (Zuckerbrot et al., 2018). A brief symptom measure characterizes current difficulties, whereas a family history measure describes a broader context of familial risk, encompassing genetic and environmental influences. Evaluating the two measures together can address two questions: whether they converge on the same young people, and whether readily obtained family history information adds to the symptom measure in identifying those with clinically significant difficulties.

The Very Quick Inventory of Depressive Symptomatology (VQIDS) and Family Index of Risk for Mood (FIRM) offer brief approaches to these complementary assessment targets. The five-item VQIDS was developed from the Quick Inventory of Depressive Symptomatology to assess core depressive symptoms (De La Garza et al., 2017), with subsequent work examining brief versions in children and adolescents (Croarkin et al., 2024). The original VQIDS covers sad mood, psychomotor retardation, negative self-outlook, reduced involvement, and low energy, with its psychometric properties extensively documented (Croarkin et al., 2024; Rush et al., 2003). A recent modification replaced psychomotor retardation with impaired concentration/decision making, because concentration was shown to be more sensitive to treatment effects in adults (Rush et al., 2026); performance of the revised VQIDS in youth is unknown. On the other hand, the FIRM uses a half-page grid to record psychiatric problems across classes of relatives, supplemented by two questions about the respondent’s own diagnoses. In its initial validation among 273 clinic referred youth, the FIRM was readily completed by parents and showed modest but significant discrimination of both pediatric bipolar disorder and unipolar depression (Algorta et al., 2013). Whether these findings extend to a national community sample and youth informant remains unclear.

It should be noted that VQIDS and FIRM require different forms of evaluation (Edwards & Bagozzi, 2000). The VQIDS treats item responses as indicators of a common symptom dimension; interpreting its total thus requires evidence concerning dimensionality, reliability, and item functioning (American Educational Research Association et al., 2014; Reise et al., 2010). In community samples where many respondents report few or no symptoms, a single reliability coefficient may obscure differences in precision across the symptom continuum. Item response theory (IRT) can clarify whether precision is concentrated in the elevated range most relevant to screening. National age- and sex-based norms would further support interpretation by situating raw scores to peers, provided that items function comparably across demographic groups and the norming models accommodate the substantial proportion of low scores. These percentiles would indicate relative symptom elevation and a potential need for further assessment.

By contrast, the FIRM aggregates reported histories across distinct problems and relatives. These indicators need not covary, so internal consistency is not an appropriate criterion for evaluating the index (Bollen & Lennox, 1991; Cicchetti et al., 2006; Streiner, 2003). Its utility instead depends on reproducible reporting, interpretable scoring, and associations with relevant outcomes. Community reference distributions can characterize the frequency and breadth of reported family psychiatric history beyond the clinical settings in which the FIRM was initially validated. Additional questions concern whether alternative scoring schemes improve on the sample total (Milne et al., 2008) and whether adolescents can report family history consistently. In the absence of diagnostic interviews, age- and sex-normed VQIDS scores provide a relevant benchmark for examining whether greater reported familial risk is associated with elevated depressive symptoms.

Using 2,252 parent reports and 1,008 linked adolescent self-reports from the BENCHMARK 2025 national survey, we first evaluated the VQIDS measurement structure, reliability, item functioning, and differential item functioning across age and sex. We then developed informant-specific age- and sex-based norms and examined parent-adolescent agreement, two-week stability, and reliable-change thresholds. This two-week period was chosen because it is roughly the gap between outpatient appointments and therefore close to the interval over which a clinician would repeat a screen. For the FIRM, we characterized score distributions, compared scoring approaches, and evaluated cross-informant agreement and retest stability. Finally, we examined associations with VQIDS percentiles, discrimination of elevated symptom benchmarks within and across informants, and convergent and discriminant associations with other symptom measures.

## Method

### Participants and Procedure

Data came from the BENCHMARK 2025 national survey conducted via YouGov (for details, see E. Youngstrom et al., 2025). Briefly, the study included parents of children aged 6-11 years, linked parent-adolescent dyads aged 12-18 years, and additional parents of adolescents. Parents with multiple eligible children were instructed to select the child whose birthday was closest to a randomly generated calendar date. Approximately 2 weeks after completing the first survey, a subset of families were invited to complete the same measures again.

The present analyses included 2,252 parent reports and 1,008 linked adolescent self-reports. Among them, 104 adolescent*-*parent dyads and an additional 104 parents of younger children completed the retest. For the retest, parents were not reminded which child they had rated initially, so we first screened for consistency in the parent’s report of the child’s characteristics for multi-child households. The basic screen required unchanged sex and an age change of no more than one year, resulting in 181 parents and 99 adolescents. The strict screen additionally checked school grade, height, and weight, resulting in 171 parents and 90 adolescents. This screening rule was not pre-registered, as the prompt failure was not planned, so results from both screens are reported; results quoted in the text are from the strict screen.

## Measures

### Very Quick Inventory of Depressive Symptomatology (VQIDS)

The VQIDS is a short form of the 16-item QIDS (Rush et al., 1012000) designed to retain coverage of the depression construct in an ultra-brief format to reduce response burden. The VQIDS employed in this report (Rush et al., 2026)has five items covering core features of depression: sad mood, impaired concentration/decision making, negative self-outlook, reduced involvement, low energy. Each item is rated on a 0-3 scale and summed to yield a total score ranging from 0 to 15.

### Family Index of Risk for Mood (FIRM)

The FIRM (parent- and self-report; Algorta et al., 2013) is a yes/no checklist asking whether any of five problems (a suicide attempt, substance problems, psychiatric hospitalization, depression, mania/bipolar disorder) has ever occurred in each of four classes of the *child’s* relatives (grandparents, parents, aunts/uncles, siblings). Two additional questions ask about the respondent’s own history of bipolar and depression diagnosis; that is, parent for the parent-report version and child for the self-report version. Each endorsed problem-by-relative cell contributes one point to the total sum score. The primary grid includes the 20 questions shared by parent- and self-report versions and ranges from 0 to 20; the expanded parent grid includes the full 22 questions and ranges from 0 to 22. As noted, FIRM items are not expected to correlate with one another (an uncle’s hospitalization needs not go with a sibling’s depression), so internal-consistency coefficients such as alpha are not meaningful (Bollen & Lennox, 1991; Cicchetti et al., 2006; Streiner, 2003).

### FIRM Comparison Measures

Parents completed: (a) the Pediatric Symptom Checklist (PSC-17; (Murphy et al., 2016)) to assess the child’s internalizing and externalizing symptoms, (b) the Child Mania Rating Scale (CMRS-21; (Pavuluri et al., 2006)) to assess mania symptoms, and (c) the Vanderbilt ADHD Diagnostic Rating Scale (VADRS; (Wolraich et al., 2003)) to assess ADHD symptoms. The two mood-related measures (PSC-17 internalizing subscale and CMRS-21) serve as convergent comparisons and the two externalizing measures (VADRS and PSC-17 externalizing subscale) as discriminant comparisons.

## Statistical Analyses

VQIDS psychometric properties were evaluated in the following order. As using sum scores assumes unidimensional factor structure, the unidimensionality of the VQIDS was first verified through confirmatory factor analysis using polychoric correlation and robust weighted least squares mean- and variance-adjusted (WLSMV) estimator. We also examined overall reliability and, importantly, how measurement precision varied across the depression continuum (Embretson & Reise, 2013; Green et al., 1984; McDonald, 2013; Zumbo et al., 2007), followed by item-level functioning using an Item Response Theory model (Kang & Chen, 2008; Orlando & Thissen, 2000; Samejima, 1968; Yen, 1984). We also tested Differential Item Functioning across sex (male vs. female) and age groups (ages 6-11 vs. 12-18) to determine whether items functioned systematically differently across groups (Meade, 2010; Swaminathan & Rogers, 1990). Next, we evaluated test-retest stability across the two-week reassessment interval (McGraw & Wong, 1996; Shrout & Fleiss, 1979) using Pearson’s *r*, Spearman’s rho, and Intraclass Correlation Coefficient (ICC; Bartko, 1966). Finally, we developed the age- and sex-based norms separately for parent- and self-report through Generalized Additive Models for Location, Scale, and Shape (GAMLSS; Rigby & Stasinopoulos, 2005).

For the FIRM, matching its formative nature, the plan was descriptive and criterion-focused rather than psychometric (Bollen & Lennox, 1991). We evaluated its national reference distribution (totals, endorsement prevalence, breadth of loading), compared alternative scoring schemes against the simple total, and quantified parent-youth agreement through ICC and item-level kappa (Cohen, 1960; Shrout & Fleiss, 1979). Whether family loading tracked the normed VQIDS benchmark was tested using a two-informant design to distinguish associations across informants from those potentially inflated by shared method variance when the same person completed both measures (Podsakoff et al., 2003; E. A. Youngstrom et al., 2015). Finally, we examined its convergent and discriminant validity against other parent-reported mood and externalizing measures (Campbell & Fiske, 1959).

## Results

### VQIDS Measurement Properties

#### Dimensionality and Internal Consistency

The five VQIDS items behaved as a single dimension: a one-factor confirmatory factor analysis model fit the data well for both parent-report data (CFI=.99, TLI=.98, RMSEA=.06, SRMR=.03) and adolescent self-report data (CFI=.98, TLI=.97, RMSEA=.09, SRMR=.04), as indicated by acceptable or good CFA fit indices (see Table 1). Internal consistency was good, with omega=.86 for parents and .88 for adolescents. The items also held together as a set without redundancy: the average correlation between items was 0.55 for parent report and 0.60 for adolescent self-report, with individual item pairs ranging from 0.47 to 0.70.

**Table 1.** Dimensionality and reliability.

|  | Parent report | Adolescent self-report |
| --- | --- | --- |
| CFI | 0.99 | 0.98 |
| TLI | 0.98 | 0.97 |
| RMSEA | 0.06 | 0.09 |
| SRMR | 0.03 | 0.04 |
| Cronbach's alpha | 0.86 | 0.88 |
| Omega | 0.86 | 0.88 |
| Marginal reliability | 0.67 | 0.73 |

*Note.* Fit indices from a single-factor WLSMV model; Cronbach's alpha and omega from the polychoric matrix; marginal reliability from the graded response model.

#### IRT Precision and Item Functioning

Test information and conditional reliability based on graded response models (Samejima, 1968) showed that the VQIDS distinguished well among children with elevated symptoms but carried little information at the low, asymptomatic end (Figure 1), supporting its use as a screen for elevated symptoms. Information was near zero below θ=−2, rose sharply as θ approaches 0, and reached a peak when θ=2, before declining at the most elevated scores (θ=4). Conditional reliability followed the same pattern, exceeding .75 above θ=0 and reaching a maximum around .85 near θ=2, then falling to .70 at θ=4. Each item’s properties are summarized in Table 2. All five items discriminated strongly, with slope (a) parameters ranging 1.50-2.33 for parent report and 1.94-2.57 for youth self-report, meaning that small differences in a child’s underlying depression level translated into clear differences in item responses. Item’s response thresholds were ordered and located in the above-average range, again reflecting VQIDS’s design as a screener. Model check was satisfactory: RMSEA values for item fit were .00-.03 across all ten item-by-informant combinations, indicating negligible misfit. The VQIDS items fit the graded response model well and showed no redundancy (no two items carried the same information over and above the total).

**Figure 1.**
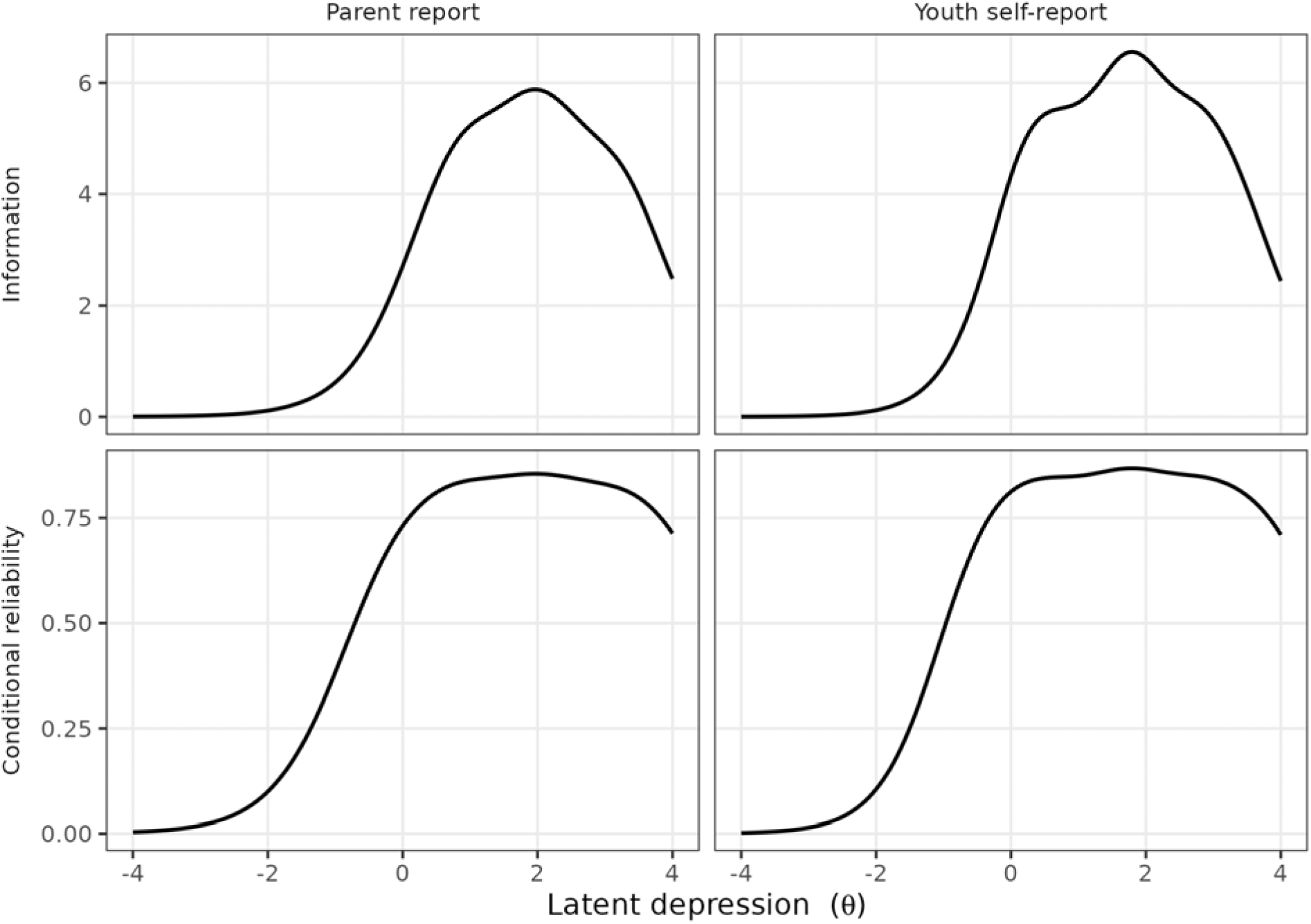
Where the VQIDS measures precisely. Test information (top) and reliability (bottom) as a function of the underlying depression level; both peak in the above-average range.

**Table 2.**
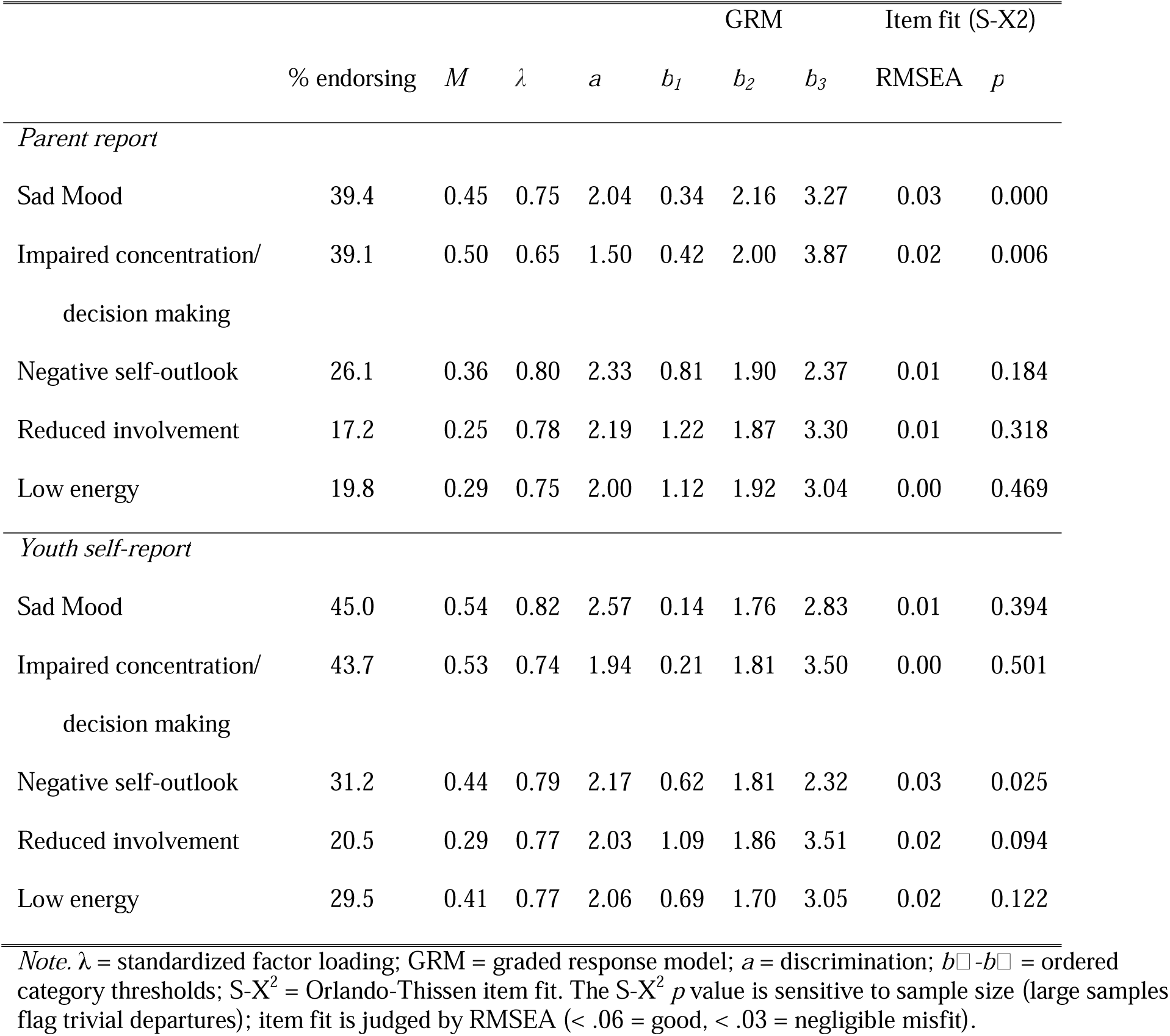
VQIDS item properties and fit.

#### Differential Item Functioning

Differential item functioning (DIF) evaluated whether any item responses differed by sex (boys vs. girls) or age (6-11 vs. 12-18 years), after accounting for the child’s actual symptom level. To avoid over-reacting to trivial differences that large samples can make “statistically significant”, we used two complementary detection methods: ordinal-logistic-regression purification (*lordif*) (Choi et al., 2011) and IRT likelihood-ratio testing (*mirt*)(Chalmers, 2012). Where the two disagreed, an item counted as DIF only if its raw expected-score difference reached 0.2 points on the 0–15 total.

The VQIDS total was essentially equivalent across all these groupings. One detection method flagged no items at all. The other flagged a couple of items on statistical grounds, but in every case the practical size of the difference was far below the 0.2-point threshold, and where two items were involved, they leaned in opposite directions and cancelled out at the total. The full per-item statistics are provided in the supplement (Tables S1-S4).

One pattern is worth noting even though it is too small to affect scoring: younger children were rated slightly higher on the low energy item, and this same small tendency appeared when adolescents rated themselves. This hints at a genuine developmental nuance rather than differences in parental reporting (Figure S1). Nevertheless, it does not change how the total should be used.

### VQIDS Parent-Adolescent Agreement

For all parent-youth dyads, we compared their VQIDS totals (Table 3) to examine cross-informant agreement. Overall agreement was moderate (ICC=.63; rank correlation about .61), with adolescents reporting slightly more symptoms than their parents, a difference that grew larger among more symptomatic children (Figure 2).

**Figure 2.**
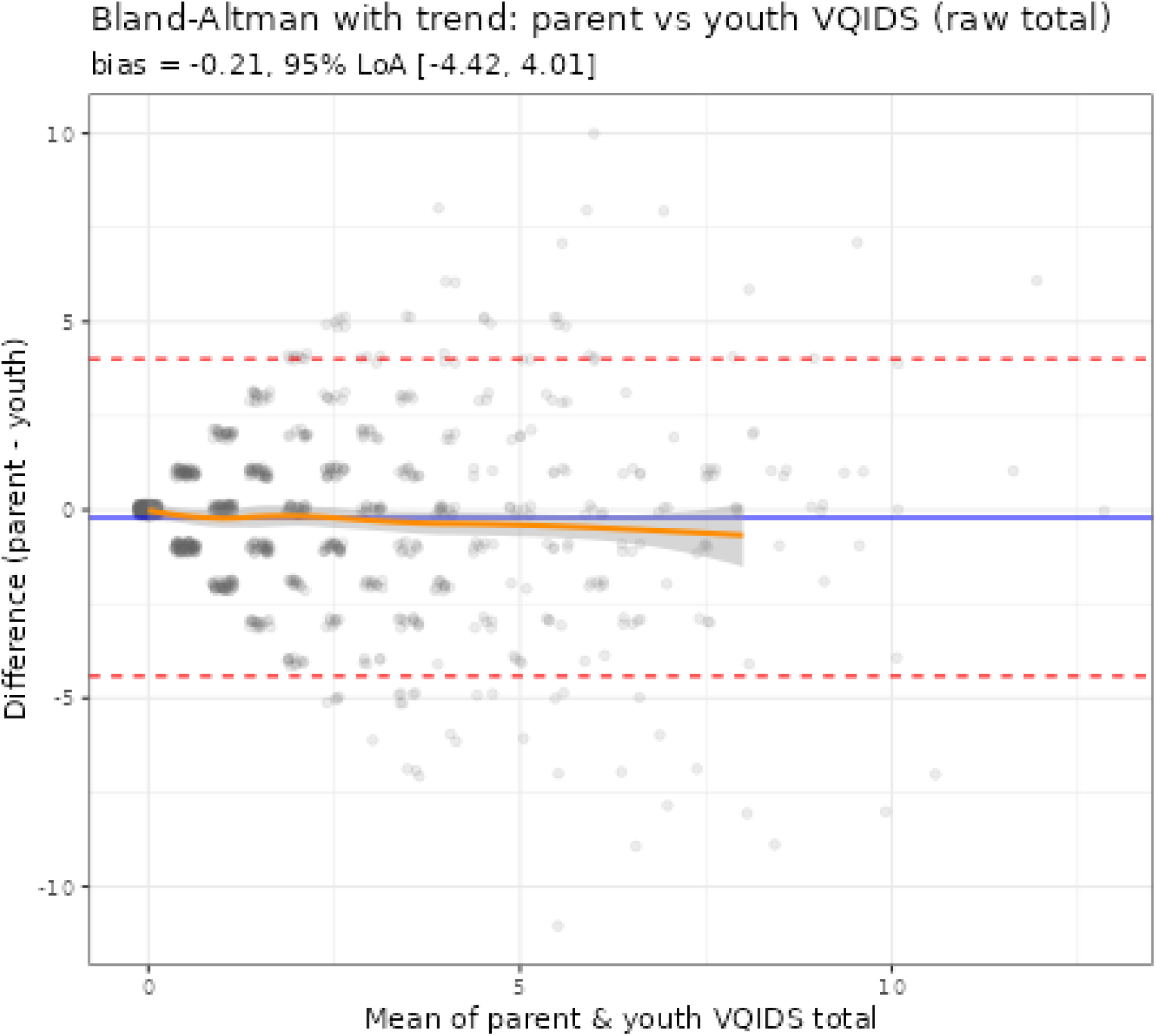
Bland-Altman plot comparing parent-youth agreement on the VQIDS (N = 1008)

**Table 3.** Parent-youth agreement, overall and among more symptomatic dyads.

|  | *N* | Rank *r* | ICC | Mean diff (P-Y) |
| --- | --- | --- | --- | --- |
| All dyads | 1008 | 0.61 | 0.63 | -0.21 |
| Drop double-zero | 783 | 0.44 | 0.54 | -0.26 |
| Both > 0 | 525 | 0.54 | 0.55 | -0.21 |
| Dyad mean $\geq 2$ | 432 | 0.22 | 0.31 | -0.33 |
| Dyad mean $\geq 3$ | 288 | 0.12 | 0.17 | -0.44 |

*Note:* ICC = intraclass correlation (absolute agreement). A positive mean difference means the parent reported more. Agreement is concentrated at the low-symptom end and declines among symptomatic dyads.

Crucially, this moderate agreement might be driven by the many families in which *both* informants report few or no symptoms: 22% of pairs were “double zeros”. Once these are set aside, agreement drops, and it continues to fall among children with elevated symptoms, precisely the range where a screen matter most. Conditioning on symptom level also mechanically narrows the range and lowers correlations, so the symptomatic-subset figures are lower bounds; but the steady decline, together with the widening parent-youth gap, points to genuine divergence rather than a statistical artifact. The practical implication is that parent and youth reports are not interchangeable for symptomatic adolescents, and it is defensible to ask the adolescent directly.

#### Two-Week Test-Retest Stability

The agreement observed between test and re-test administrations was moderate: Pearson’s *r*=.61 for parent-report and *r*=.56 for adolescent self-report, with ICC of .59 and .55, respectively (Table 4). Results for sample using basic screen are comparable.

**Table 4.** VQIDS total across a two-week retest interval.

|  | Baseline |  |  | Recontact |  | Mean change | ICC |  |  |
| --- | --- | --- | --- | --- | --- | --- | --- | --- | --- |
| | *n* | *M* | *SD* | *M* | *SD* | [95% *CI*] | *r* | $\rho$ | [95% *CI*] |
| Strict same-child screen |  |  |  |  |  |  |  |  |  |
| Parent report | 171 | 1.99 | 2.25 | 1.53 | 1.90 | -0.47 [-0.75, -0.19] | 0.61 | 0.61 | 0.59 [0.50, 0.69] |
| Youth self-report | 90 | 2.38 | 2.66 | 1.91 | 2.48 | -0.47 [-0.97, 0.04] | 0.56 | 0.45 | 0.55 [0.39, 0.68] |
| Basic same-child screen. |  |  |  |  |  |  |  |  |  |
| Parent report | 181 | 1.99 | 2.25 | 1.50 | 1.92 | -0.49 [-0.75, -0.22] | 0.63 | 0.62 | 0.61 [0.52, 0.70] |
| Youth self-report | 99 | 2.45 | 2.63 | 2.08 | 2.57 | -0.37 [-0.84, 0.10] | 0.59 | 0.51 | 0.59 [0.44, 0.70] |

*Note.* *r* = correlation between occasions; $\rho$ = rank correlation; ICC = intraclass correlation ICC(2,1), absolute agreement, which unlike correlation is sensitive to mean-level drift. Change in recontact minus baseline. The two screens compare each family's two sittings on the child's sex, age, school grade, height, and weight to guard against a parent reporting on a different child at recontact; both are shown because the rule was not pre-registered.

On average, scores fell by about half a point at retest for both informants (−0.47 for parents, −0.47 for adolescents). For adolescents only, the rank correlation (.45) was lower than the ordinary correlation (.56), suggesting that the linear association may be disproportionately influenced by a small number of high-scoring adolescents, with weaker correspondence among those scoring near the floor.

#### Reliable Change

To evaluate whether a change between two visits is large enough to act on, we take into consideration the spread of scores in the population and a reliability coefficient. From these we derive the standard error of measurement attached to a single score, then the standard error of a difference between two scores, and finally the change large enough to it would be exceeded by chance only 10% or 5% of the time (Jacobson & Truax, 1991). Alongside these we report a minimally important difference, approximately as half a standard deviation.

We report Jacobson and Truax’s original deviation using test-retest dependability, which is the coefficient that describes error accumulating across occasions, and, reliable-change, that always substitutes an internal-consistency coefficient. The latter describes error arising from particular items included in the scale (out of the universe of possible depression items), but it does not capture sources of variability that arise between one assessment and the next. These two coefficients therefore are not interchangeable, and, for this reason, both are reported. For parent report, the 95% reliable change threshold was 2.4 points on the internal-consistency coefficient and 4.0 points on the retest coefficient; for youth self-report, the corresponding thresholds were 2.5 and 4.7 (see Table 5).

**Table 5.** How much a VQIDS total must change before it exceeds measurement error.

|  | *SD* | Reliability | *SEM* | *SE<sub>diff</sub>* | Reliable change |  |  |
| --- | --- | --- | --- | --- | --- | --- | --- |
|  |  |  |  |  | 90% | 95% | *MID* |
| Parent report |  |  |  |  |  |  |  |
| Internal consistency (omega) | 2.32 | 0.86 | 0.87 | 1.23 | 2.02 | 2.40 | 1.16 |
| Retest dependability | 2.32 | 0.61 | 1.44 | 2.04 | 3.36 | 4.00 | 1.16 |
| Adolescent self-report |  |  |  |  |  |  |  |
| Internal consistency (omega) | 2.56 | 0.88 | 0.88 | 1.25 | 2.06 | 2.45 | 1.28 |
| Retest dependability | 2.56 | 0.56 | 1.71 | 2.41 | 3.97 | 4.73 | 1.28 |

*Note.* $SEM = SD \times \sqrt{1 - \text{reliability}}$ ; $SE_{diff} = SEM \times \sqrt{2}$ . A raw-score change of at least the tabled value is unlikely to be measurement error alone (Jacobson & Truax, 1991). *MID* = minimally important difference, approximated as half a standard deviation. *SD* and internal-consistency reliability are from the full baseline sample; the retest row substitutes the observed dependability coefficient for omega.

From a practical point of view, on a scale whose standard deviation is about 2.3 points, the wider threshold means a child must move roughly 1.7 standard deviations before the change can be called real: a child scoring 5 at the first visit would need to fall to 1 or below at the second.

### VQIDS Age- and Sex-Based Norms

VQIDS totals are strongly skewed toward the low end, as expected for a depression screen in a general population: about 38% of parent reports and 33% of youth self-reports were exactly 0, and most of the remaining scores were low (Table 6). This “floor” means that elevated scores are genuinely uncommon and therefore informative, indicating that the norms must be built with a model suited to such data distribution.

**Table 6.** Distribution of VQIDS totals.

|  | *N* | *M* | *SD* | Median | % scoring 0 | Skew |
| --- | --- | --- | --- | --- | --- | --- |
| Parent report | 2252 | 1.85 | 2.32 | 1 | 37.9 | 1.65 |
| Adolescent self-report | 1008 | 2.21 | 2.56 | 1 | 32.6 | 1.50 |

*Note.* Totals range 0 to 15. High skew and a large share of zero scores reflect the expected floor of a depression screen in a general-population sample.

In the GAMLSS specification, we modelled the score distribution as a function of age and sex separately for each informant, allowing the typical score and its spread to change smoothly with age. The distribution itself was not assumed (Micceri, T., 1989): we compared three candidate count distributions (R. Rigby et al., 2017) and retained the Negative Binomial distribution as the best fitting one, with the comparison and its diagnostics reported in Supplement S7. From this GAMLSS model, any raw VQIDS total can be expressed as a percentile for a child of a given age and sex. For example, “a score of 3 is around the 85^th^ percentile for a 14-year-old girl.” Because the raw scores are whole numbers with a large floor, each score spans a small band of percentiles; we report the midpoint of that band, which gives a single, reproducible value. Two practical points follow. First, because each child is compared only to same-age, same-sex peers, a percentile-based cutoff automatically applies an even standard across boys and girls and across ages. Second, since scores come in lumps, a “top 10%” flag will not capture exactly 10% of children at every age (it may be a little more), which is a property of the data and is reported transparently in the tables.

Because many clinicians read *T-*scores by habit, each percentile is also reported as a normalized *T-*score: *T =* 50 +10*z*, where z is the normal deviate of that same age-and sex-conditional percentile. Carrying the exact same information as the percentile does, *T-*score is a relabelling for familiarity, and a given percentile maps the same T at every age and sex (the 70^th^ percentile is *T =* 55, the 80^th^ *T =* 58, the 90^th^ *T =* 63, and the 95^th^ *T =* 66). We did not derive T from the item response theory score used in Supplement S6, because a theta is calibrated on the pooled sample and is not conditioned on age or sex; converting it would quietly average over the developmental norming.

The zero-compiled floor displayed in Table 5 affects the *T-*scale such that it is easy to misread: A raw score of 0 does not sit at the bottom of the *T* distribution. If falls at roughly the 17 to 20^th^ percentile for parent report (*T =* 40 to 42) and the 13 to 18^th^ for youth self-report (*T =* 39 to 41), which is the middle-low range rather than the extreme. This is ordinary for a one-sided screener and follows from how the scale has been framed throughout: it is built to flag elevation, not to grade how well a child is doing at the low end. Here a low *T* means “nothing flagged”, not “unusually good mood.”

Table 7 summarizes, for each reporter, the lowest raw score that reaches key percentile benchmarks (80^th^, 90^th^, 95^th^) at representative ages; because the scale floor heavy, these cutoffs are low and vary little across age. The complete single-year lookup tables (raw score to percentile and *T-*score, and percentile-to-cutoff, for each reporter) are provided in the Appendix (Tables A1-A4) for clinical use.

**Table 7.**
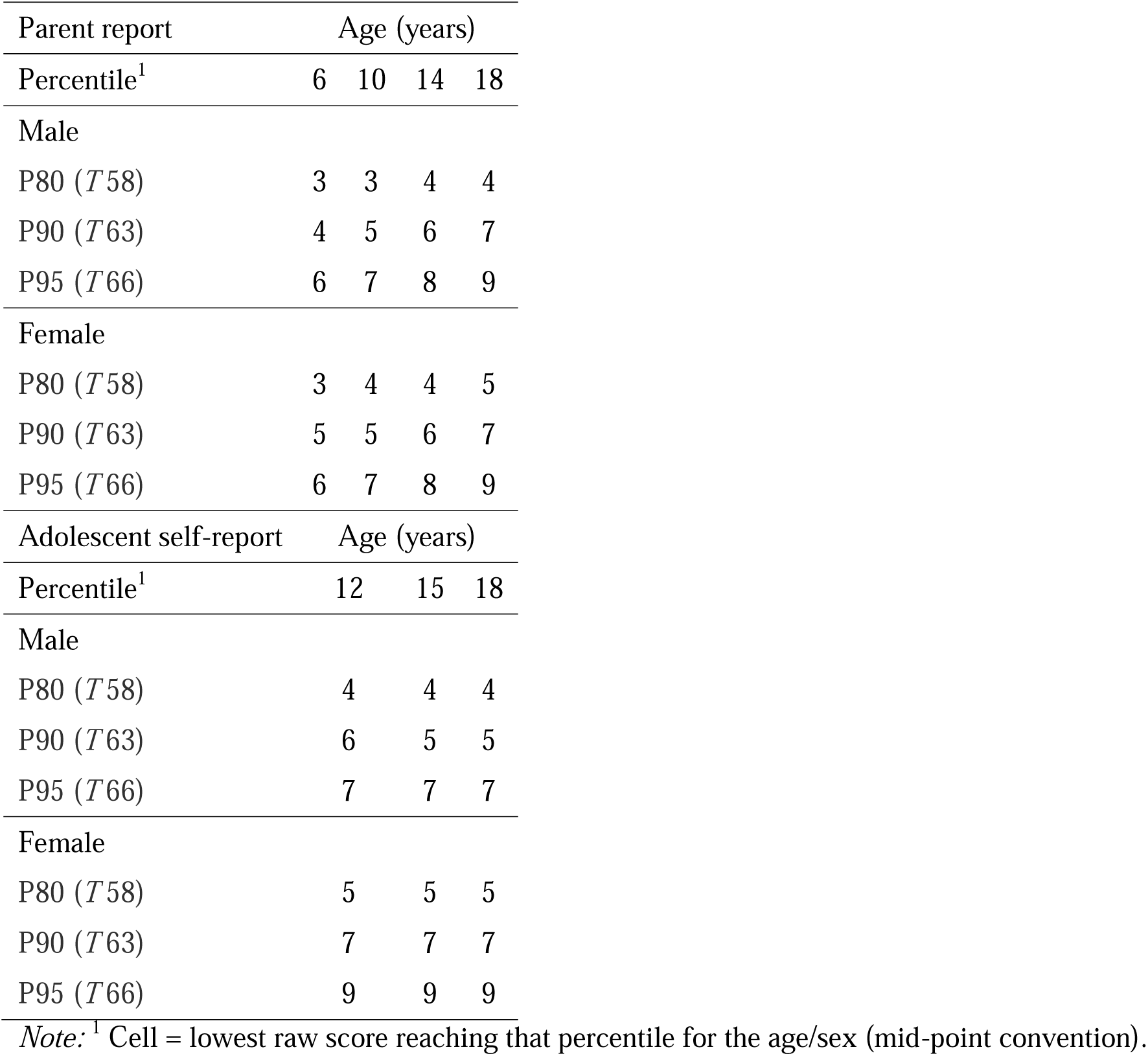
Parent report and Youth self-report lowest raw score reaching each percentile, at representative ages.

### FIRM Reference Distributions

Family mental history was common but usually limited in scope: 69% of parents endorsed at least one of the 20 grid items, yet the median grid total was 2 of 20 and the distribution was strongly right-skewed (Table 8). The typical family reported loading in 1.8 of the five problem areas, spread across 1.4 of the four relative classes. On the two self-report items, 9.9% of caregivers reported having been diagnosed with bipolar disorder themselves and 39.7% with depression.

**Table 8.** Distribution of FIRM totals (Parent report)

|  | *N* | *M* | *SD* | Median | % scoring 0 | Skew |
| --- | --- | --- | --- | --- | --- | --- |
| Primary grid total (0–20) | 2252 | 2.69 | 2.92 | 2 | 31.0 | 1.28 |
| Parent expanded grid total (0–22) | 2252 | 3.18 | 3.29 | 2 | 27.3 | 1.19 |

*Note:* The grid total sums the 20 relative-grid items (primary score); the full total adds the two caregiver self-report items (own bipolar and depression diagnoses).

Endorsement was not uniform across content (Table 9): depression was the most reported problem area (57% of families endorsed it for at least one relative class) and suicide attempt the least (21%). By relative class, loading was reported most often for parents (47% and least often for siblings (13%).

**Table 9.** Family-history endorsement by problem area and by relative class (parent report)

|  | % of families endorsing | Mean items endorsed |
| --- | --- | --- |
| By problem area (any of 4 relative classes) |  |  |
| Suicide attempt | 21.4 | 0.27 |
| Substance problems | 49.2 | 0.76 |
| Psychiatric hospitalization | 23.6 | 0.29 |
| Depression | 57.4 | 0.99 |
| Mania/bipolar | 29.0 | 0.38 |
| By relative class (any of 5 problem areas) |  |  |
| Grandparents | 41.5 | 0.78 |
| Parents | 47.0 | 0.93 |
| Aunts/uncles | 38.7 | 0.73 |
| Siblings | 13.3 | 0.24 |

*Note:* Problem-area counts range 0–4 (one item per relative class); relative-class counts range 0–5 (one item per problem area).

### FIRM Alternative Scoring Approaches

A formative index like the FIRM invites alternative scoring rules: count only mood problems, only bipolar, weight first-degree relatives more heavily (parents and siblings share twice the proportion of genes that grandparents and aunts or uncles do), or simply count how many relative classes are affected at all. Table 10 compare main candidates.

**Table 10.** Rank correlations among alternative FIRM scoring schemes and the VQIDS percentile (parent report)

|  | 1 | 2 | 3 | 4 | 5 | 6 |
| --- | --- | --- | --- | --- | --- | --- |
| 1. Grid total (0-20) |  |  |  |  |  |  |
| 2. Full total (0-22) | 0.98 |  |  |  |  |  |
| 3. Mood-only (suicide + depression + bipolar) | 0.94 | 0.93 |  |  |  |  |
| 4. Bipolar-only | 0.66 | 0.65 | 0.69 |  |  |  |
| 5. Breadth (relative classes affected) | 0.93 | 0.91 | 0.87 | 0.53 |  |  |
| 6. Weighted first-degree | 0.99 | 0.97 | 0.93 | 0.65 | 0.92 |  |
| 7. VQIDS percentile | 0.27 | 0.29 | 0.29 | 0.21 | 0.25 | 0.27 |

*Note:* Spearman correlations. Mood-only sums the suicide, depression, and bipolar domains. Weighted first-degree counts parents and siblings twice: these first-degree relatives share, on average, twice the proportion of genes that the second-degree grandparents and aunts/uncles do, so the 2:1 weighting mirrors the kinship coefficients (.50 vs .25). Breadth counts how many relative classes have any endorsement.

The different scoring strategies were strongly correlated (rank correlations .53 to .99), which is to say they largely re-describe the same family loading. Their associations with the VQIDS percentile likewise fell in a narrow band, from .21 to .29. The simple grid total sat in the middle of that band (.27); the mood-only and full totals edged it by a rounding margin (both .29), and the narrowest scheme, bipolar-only, was the weakest (.21). Restricting to count to mood problems, or weighting relatives by closeness, therefore buys nothing worth the added complexity, echoing the original validation study (Algorta et al., 2013). The grid total is retained as the primary score. The one comparison that is worth a formal test, the grid total against the full total (which adds the caregiver’s own diagnoses), is made in the discrimination analyses below, where the two can be tested on the same children against the same benchmark.

### FIRM Parent-Adolescent Agreement

Cross-informant agreement on the primary grid total was good: ICC(2,1) = .74 [.71, .77] (Figure 3), rank correlation .74, with parents reporting slightly more than adolescents on average (difference 0.24 points, *p*<.001). This is visibly stronger than the agreement on the VQIDS symptom totals in the same dyads (Table 7), the ordering one would predict: family history is closer to a matter of fact, whereas symptom report is a perception.

**Figure 3.**
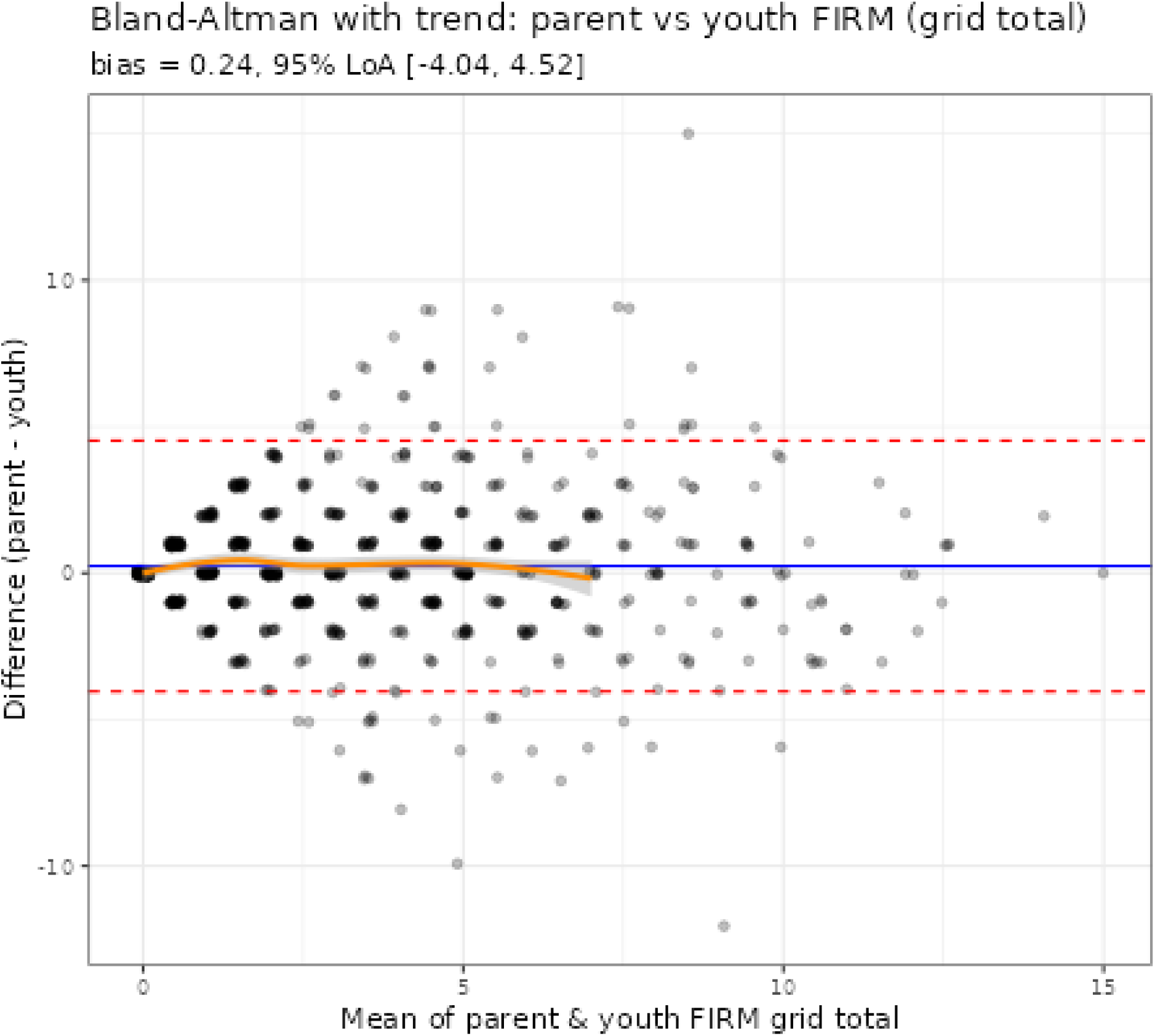
Bland-Altman plot comparing parent-youth agreement on the FIRM (N = 1008)

Agreement also tracked how knowable the information is (Table 11). It was higher for closer relatives (parents: ICC=.74; grandparents: .69; siblings: .66; aunts/uncles: .61) and higher for the problems a family is more likely to know about and speak about openly. A relative’s depression, which tends to be enduring, acknowledged, and often treated, drew closer agreement (ICC=.67) than a suicide attempt (.54), which is a single event, heavily stigmatized, and one that adults often deliberately keep from children. That gradient is itself a validity signal: the two informants converge most where the fact is easiest to learn, as one would expect if both are reporting on a real family history rather than answering out of habit or style. One caveat belongs with it: suicide attempts are also rarer in these families than depression (21% versus 57% of families), and agreement statistics are mechanically impacted for rare events, so part of this gap may be base rate rather than concealment. Item-level agreement for the 20 individual grid items is given in the supplement (Table S10); kappas ranged from .39 to .69, and should be read alongside each item’s prevalence, since kappa is affected for rare facts even when raw agreement is high.

**Table 11.** Parent-adolescent agreement on the FIRM.

|  | **Parent *M*** | **Adolescent *M*** | **Rank *r*** | **ICC** |
| --- | --- | --- | --- | --- |
| Primary grid total (0-20) | 2.99 | 2.75 | 0.74 | 0.74 |
| Problem: Suicide attempt | 0.26 | 0.25 | 0.56 | 0.54 |
| Problem: Substance problems | 0.86 | 0.77 | 0.64 | 0.65 |
| Problem: Psychiatric hospitalization | 0.34 | 0.34 | 0.63 | 0.64 |
| Problem: Depression | 1.12 | 1.01 | 0.68 | 0.67 |
| Problem: Mania/bipolar | 0.42 | 0.38 | 0.65 | 0.62 |
| Relative class: Grandparents | 0.81 | 0.67 | 0.66 | 0.69 |
| Relative class: Parents | 1.05 | 1.02 | 0.74 | 0.74 |
| Relative class: Aunts/uncles | 0.82 | 0.73 | 0.59 | 0.61 |
| Relative class: Siblings | 0.32 | 0.33 | 0.61 | 0.66 |
| Breadth: relative classes affected | 1.55 | 1.43 | 0.68 | 0.68 |
| Breadth: problem areas affected | 1.94 | 1.80 | 0.72 | 0.73 |

*Note:* $N = 1008$ dyads. ICC = intraclass correlation ICC(2,1), absolute agreement. Problem-area counts range 0–4, relative-class counts 0–5. Both informants describe the same family; the two caregiver self-report items are excluded because they refer to the reporter's own diagnosis.

### Two-Week Test-Retest Stability

The re-test data provided an opportunity to evaluate the reliability of the FIRM. Agreement across two weeks was good, and clearly better than the VQIDS symptom scale: Pearson’s *r*=.80 for parents and .76 for adolescents, with ICC of .79 and .75 (Table 12).

**Table 12.** FIRM grid total across a two-week interval.

|  | n | Baseline<br>M | Baseline<br>SD | Recontact<br>M | Recontact<br>SD | Mean change<br>[95% CI] | r | ρ | ICC<br>[95% CI] |
| --- | --- | --- | --- | --- | --- | --- | --- | --- | --- |
| **Strict same-child screen** |  |  |  |  |  |  |  |  |  |
| Parent report | 171 | 2.88 | 3.13 | 3.12 | 3.44 | 0.24 [−0.08, 0.56] | 0.80 | 0.79 | 0.79 [0.73, 0.84] |
| Adolescent self-report | 90 | 2.80 | 2.82 | 3.06 | 3.33 | 0.26 [−0.20, 0.71] | 0.76 | 0.75 | 0.75 [0.65, 0.83] |
| **Basic same-child screen** |  |  |  |  |  |  |  |  |  |
| Parent report | 181 | 2.81 | 3.09 | 3.05 | 3.41 | 0.24 [−0.06, 0.55] | 0.80 | 0.79 | 0.79 [0.73, 0.84] |
| Adolescent self-report | 99 | 2.86 | 2.82 | 3.05 | 3.25 | 0.19 [−0.24, 0.62] | 0.76 | 0.75 | 0.75 [0.65, 0.83] |

*Note:* $r$ = correlation between occasions; $\rho$ = rank correlation; ICC = intraclass correlation ICC(2,1), absolute agreement. Change is recontact minus baseline. Grid total (0–20); the two caregiver self-report items are excluded. Screens as in Table 2.

It is worth being clear about what these figures do not show. Family history cannot change in two weeks. A grandparent’s suicide attempt either happened or didn’t happen, so agreement here should in principle be 1.0. At an intraclass correlation of .79, roughly 36% of the variance in reported family loading belongs to the occasion of asking rather than the family itself. Therefore, family history carries more occasion-to-occasion error than its factual content would suggest, which argues for treating a single administration as an estimate rather than a record, in the same spirit as the item-level caution above. Reporting loading also rose slightly at recontact for both reporters (0.24 and 0.26 points), consistent with recall improving on a second asking, though neither difference is distinguishable from zero.

#### Associations with Depressive Symptoms

FIRM criterion associations were evaluated against age- and sex-normed VQIDS benchmark. Because diagnostic interviews were unavailable, these analyses concern depressive-symptom elevation rather than diagnostic accuracy.

##### Symptom Percentiles

The FIRM grid total correlated with the VQIDS percentile at rho=.27 for parent report (Kruskal-Wallis across percentiles bands *p*<.001) and rho =.36 for adolescent self-report (*p*<.001; Figure 2). The association is modest in absolute terms, as expected for a distal risk factor, but it is graded.

##### Discrimination of Elevated Symptom Scores

Table 13 reports all FIRM-informant by VQIDS-informant combinations in the same dyads using the 90^th^ percentile cutoff. Same-informant associations may include shared-rater variance and should be treated as upper bounds; cross-informant associations examine whether discrimination persists when different people provide the predictor and criterion.

**Table 13.** FIRM discrimination of the 90^th^-percentile VQIDS benchmark: the two-informant matrix.

|  | Criterion |  |
| --- | --- | --- |
| | Parent VQIDS $\geq 90^{\text{th}}$ | Adolescent VQIDS $\geq 90^{\text{th}}$ |
| Parent FIRM, primary grid total | .677 [.624, .731] | .670 [.617, .724] |
| Parent FIRM, expanded grid total | .691 [.639, .742] | .691 [.640, .742] |
| Adolescent FIRM, grid total | .698 [.647, .749] | .731 [.682, .780] |

*Note:* AUROC [95% CI]; 0.5 = chance. All cells computed on the same 1008 dyads classifiable on both benchmarks. Cells where predictor and criterion share a reporter (Parent FIRM rows under the parent criterion; Adolescent FIRM rows under the adolescent criterion) include shared-rater variance and are upper bounds; the cross-reporter cells are the method-clean tests. The full total appears for parent report only: the adolescent form's two extra items ask about the adolescent's own diagnoses rather than a relative's, so they are personal rather than family history.

Cross-informant AUROCs ranged from .670 to .698, all above chance. Shared-rater inflation was small: moving each FIRM from its own reporter’s criterion to the other reporter’s criterion only lowered .007 (parent FIRM) and .033 (adolescent FIRM) in AUROC. Holding the criterion constant to ask whose report carries more signal: against the parent’s benchmark, the adolescent’s FIRM discriminated at .698 versus .677 for the parent’s own FIRM; against the adolescent’s benchmark, .731 versus .670. Adolescents, in short, can report family history to good effect.

Adding parent’s own depression and bipolar diagnoses improved parent FIRM discrimination against the parent benchmark (.677 to .691; *p*=.033). The natural potential caveat is that this gain is circular: a parent who describes their own depression may also describe more symptoms in their child, which would inflate the improvement without any real information being added. That situation can be tested by changing the benchmark to the adolescent’s own report, a second source rather than sharing method variance (Campbell & Fiske, 1959; Podsakoff & Podsakoff, 2012), and the improvement held there as well (.670 to .691; *p*=.001). The parent’s own history therefore carries genuine information about the child’s risk, not merely an echo of how that parent answers questionnaires. Discrimination at the more lenient 70th and 80th percentile benchmarks is reported in the supplement (Table S11).

#### Convergent and discriminant validity

If the FIRM indexes mood risk, its total should be more associated with mood and internalizing symptoms than with unrelated externalizing problems. We correlated the FIRM grid total with two mood/internalizing scales (the parent-reported internalizing subscale of the Pediatric Symptom Checklist, and the Child Mania Rating Scale) and two externalizing/attention scales (the Vanderbilt ADHD rating scale and the externalizing subscale of the same checklist), all completed by the parent (Table 14; *n*=2251 parents with the comparison scales).

**Table 14.** Convergent and discriminant validity of the FIRM (parent report)

|  | Convergent (mood) |  | Discriminant (externalizing/attention) |  |
| --- | --- | --- | --- | --- |
|  | Internalizing | Mania (CMRS) | ADHD (Vanderbilt) | Externalizing |
| VQIDS symptom screen | 0.58 | 0.52 | 0.49 | 0.40 |
| FIRM primary grid total | 0.35 | 0.24 | 0.30 | 0.20 |

*Note:* Spearman correlations; $n = 2251$ parents. Internalizing and externalizing are the two subscales of the Pediatric Symptom Checklist. A mood-risk index should correlate more with the convergent (mood) scales than the discriminant (externalizing) ones. The within-checklist internalizing-versus-externalizing gap for the FIRM is statistically reliable (Steiger's test, $p < .001$ ).

The prediction was confirmed. On the symptom checklist, the FIRM correlated with internalizing symptoms (.35) more strongly than with externalizing symptoms (.20), a gap unlikely to be chance (*p*<.001). Because both come from the same checklist, that contrast cannot be explained away by differences in scale length or format. The two externalizing markers were both low, the Vanderbilt ADHD scale at .30 and the externalizing subscale at .20, well below the FIRM’s tie to internalizing symptoms.

One convergent result was weaker than expected and is worth to be mentioned here: the Child Mania Rating Scale correlated only .24 with the FIRM, in the same low range as the externalizing markers rather than with its internalizing companion. The likeliest reason is that current mania symptoms are near the floor in a general-population sample, giving a family-history index little child-level mania to track; here the FIRM’s mood signal is carried by internalizing symptoms, not mania. As a reference, the VQIDS symptom screen showed the expected ordering too (Table 14, top row), tracking the mood scales more closely than the externalizing ones.

Supplemental analyses evaluated the classification accuracy of the VQIDS and FIRM for identifying cases with a history of depression or bipolar disorder as reported by either the parent or youth. Both scales showed good accuracy for depression, with AUCs of .72 to .77 (Table S12), and bipolar, with AUCs of .73 to .83 (Table S13). These are aligned with estimates from meta-analyses. The parent and youth agreed well about the youth’s diagnoses, with Gwet’s AC values of .87 for depression and .97 for bipolar (Table S14). Logistic regression analyses found that both scales were well calibrated, and combining the scales within and across informants also significantly improved classification accuracy, consistent with evidence-based assessment recommendations (Table S15). All of these statistics are more impervious to low base rates and score floor effects than correlations are. Even so, we do not report a cut score, because the diagnoses are based on “diagnosis as usual” in the community and reported by the family rather than a professional; agreement is known to be low between these diagnoses and those based on structured diagnostic interviews (Jensen-Doss et al., 2014; Rettew et al., 2009).

## Discussion

This study evaluated two complementary components of brief youth mood assessment in a large, nationally representative sample: current depressive symptoms, assessed with the VQIDS, and familial psychiatric risk, assessed with the FIRM. The findings support distinct roles for the two measures. The VQIDS functioned as a brief, largely unidimensional screen whose precision is concentrated at elevated symptom levels, with age- and sex-based norms providing a practical framework for interpreting scores. The FIRM provided a simple summary of reported family psychiatric loading that was reasonably reproducible and associated with elevated depressive symptoms even when family history and symptoms were reported by different informants. Considered together, the measures capture different information: the VQIDS characterizes how youth is doing currently, whereas the FIRM provides context about familial vulnerability.

Several aspects of the VQIDS findings are especially important for interpretation. First, good overall internal consistency should not be taken to mean that the measure is equally precise everywhere along the symptom continuum. Its information is concentrated in the elevated range, while the substantial floor in this community sample limits differentiation among youth with few symptoms. This is appropriate for a screener designed to identify elevations rather than finely grade the low end of the distribution. The age- and sex-conditional norms make that distinction more explicit: a high percentile indicates that symptoms stand out relative to same-age, same-sex peers and may warrant closer assessment, whereas a low *T-*score should not be interpreted as unusually good mental health. The absence of practically meaningful differential item functioning across age and sex further supports use of the same total score across these groups.

Second, the VQIDS appears better suited to screening than to detecting small changes over time. Two-week stability was moderate, and using retest dependability produced relatively wide reliable-change thresholds. Thus, a movement of one or two raw-score points may be clinically interesting but cannot confidently be distinguished from occasion-to-occasion variability using this brief measure alone. Conversely, the reliable change thresholds reported here indicate when change is unlikely to reflect measurement error; they do not establish that the change is itself clinically important to the child or family. The distribution based minimally important differences should be interpreted similarly rather than as empirically validated treatment response thresholds. Services primarily interested in monitoring incremental improvement may therefore benefit from a longer measure with greater longitudinal precision. Parent and adolescent reports also should not be treated as interchangeable. Their agreement became weaker as symptoms increased, precisely where assessment decisions become more consequential. Obtaining the adolescent’s own report when feasible therefore adds information rather than simply duplicating the parent’s report.

The FIRM requires a different interpretive framework because its items describe distinct family history facts rather than interchangeable indicators of a latent trait. The present results provide little reason to replace the straightforward grid total with more elaborate weighting or content specific scores. More restrictive scoring largely reorganized the same information without producing a clear gain in association with depressive symptoms. At the same time, the findings illustrate an important limitation of treating family history as purely objective information. Parent-adolescent agreement was stronger for family history than for current symptoms and was greatest for information that should be easier for both informants to know, yet reporting was not perfectly stable even across two weeks. FIRM scores therefore reflect not only the family’s psychiatric history but also what the respondent knows, recalls, and is willing or able to report.

The cross-informant analyses strengthen the evidence that FIRM scores capture meaningful familial risk rather than simply a general tendency for one respondent to endorse problems. Greater family loading remained associated with elevated VQIDS scores when one person reported family history and the other reported symptoms, and adolescents themselves provided useful family history information. The pattern was nevertheless more consistent with a broad familial psychiatric risk indicator than with a narrowly mood specific measure. FIRM scores related most clearly to internalizing symptoms, but the association with current mania symptoms was weaker. Diagnostic and prospective data will be needed to determine how specifically the index predicts depressive or bipolar disorders.

The combined analyses further clarify why symptom screening and family history may be useful together. Models containing both sources of information generally discriminated the reported diagnostic outcomes better than models using either measure alone, although individual parent- or adolescent-report predictors sometimes became redundant once information from all reporters was included. This pattern is consistent with complementarity rather than interchangeability: current symptoms and familial history each contribute information, but their incremental value depends on what is already known. Because the diagnostic outcomes in this survey were themselves based on questionnaire reports rather than structured diagnostic interviews, these findings should be viewed as evidence of nonredundancy within the present assessment battery rather than evidence that combining the measures improves clinical diagnosis or patient outcomes. Still, the results are consistent with recommendations to include gathering family history as well as using validated scales to assess for possible mood disorder (Hankin & Cohen, 2020; Keramatian et al., 2023; E. A. Youngstrom, 2026).

The national community sampling is an important strength for establishing reference distributions and examining these measures outside clinically referred samples. It also provides broader demographic heterogeneity than the clinical samples in which brief mood measures and family history indices are often studied. Nevertheless, generality should not be assumed beyond what was tested. Measurement equivalence of the VQIDS was evaluated across age and sex, but not across racial, ethnic, socioeconomic, or other demographic groups, and the present norms should be replicated before applied to substantially different populations or settings. Adolescent self-report was also available only from ages 12 to 18. Thus, the national design strengthens the relevance of the findings to U.S. community screening, but does not establish equivalent performance in clinical samples, other countries, or every demographic subgroup.

Other limitations qualify the conclusion. Symptoms and family history were not verified by diagnostic interviews, and the principal associations were cross-sectional. Consequently, the study cannot determine diagnostic accuracy, predict future disorder onset, or establish whether adding FIRM to symptom screening improves clinical decisions. The smaller retest sample and uncertainty about whether parents rated the same child also constrain stability estimates, although results were similar across screening rules. FIRM totals additionally depend on family composition and informants’ opportunities to learn about relatives, neither of which is captured by a simple endorsement count. Longitudinal measurement invariance was not evaluated, so retest correlations do not establish equivalent VQIDS item functioning across occasions (Liu et al., 2017).

Replication in independent community and clinical samples should evaluate the normative thresholds against diagnoses, impairment, and longitudinal outcomes. Future work could also test whether family history information improves referral or monitoring decisions beyond symptom scores alone and whether score-specific precision improves interpretation of change. The present findings provide the measurement and descriptive foundation for those evaluations while retaining the practical advantages of brief parent and adolescent reports.

## Data Availability

All data produced in the present study are available upon reasonable request to the authors

## Author Note

Guillermo Perez Algorta, Division of Health Research, Faculty of Health and Medicine, University of Lancaster, UK; Lilian Li, Institute for Mental and Behavioral Health Research, Nationwide Children’s Hospital and Department of Psychiatry and Behavioral Health, The Ohio State University; Yinuo Liu, Institute for Mental and Behavioral Health Research, Nationwide Children’s Hospital; Eric A. Youngstrom, Institute for Mental and Behavioral Health Research, Nationwide Children’s Hospital and Department of Psychiatry and Behavioral Health, The Ohio State University; Phoebe A. Rodda, Gillings School of Global Public Health, University of North Carolina at Chapel Hill; A. John Rush, Department of Psychiatry, Duke-National University of Singapore.

## Acknowledgements

Supported by gifts from the Chlapaty, DiMarco, and Zipfel families to Nationwide Children’s Hospital to support the Institute for Mental and Behavioral Health Research (IMBHR). The authors gratefully acknowledge Jonathan Covarrubias for his assistance with the clinical dataset, and Marissa McClellan, Kevin Stephenson, Jacqui Pazaropoulos, Brittany Grate, and Kim Jones for their contributions to the development and implementation of the BENCHMARK project, including literature and reference review, study coordination and administration, data organization and management, data cleaning, and development of analytic resources.

