## Supplemental Material for "Small Enough to Use, Sharper Together: VQIDS and FIRM in U.S. Youth Ages 6–18"

**VQIDS Measurement and Norms, and FIRM Characterization**

Supplemental Figures and Tables

**Figure S1**

*Expected item score by age band and reporter for the two items with a detectable but negligible developmental gradient (Self-Outlook and Psychomotor Slowing). Differences are below the practical threshold and do not affect total-score interpretation.*

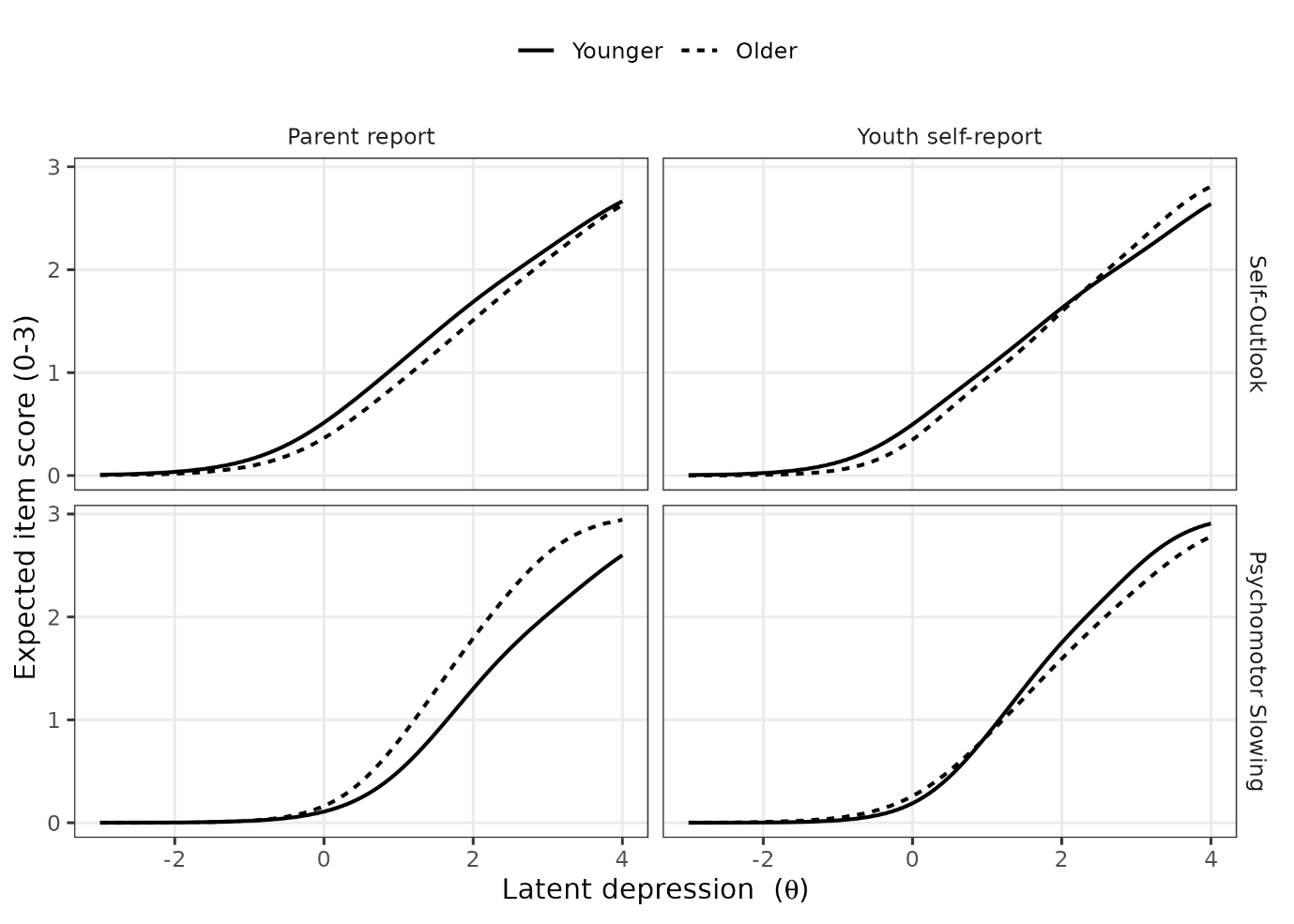

**Figure S2**

*Mean FIRM total (95% CI) across VQIDS percentile bands, by reporter. Within each panel, predictor and criterion come from the same reporter; the cross-reporter tests are in Table 13 in main paper. The full total (cyan) is shown for parent report only; on the adolescent form the two extra items ask about the adolescent’s own diagnoses rather than a relative’s, so they are not family history*
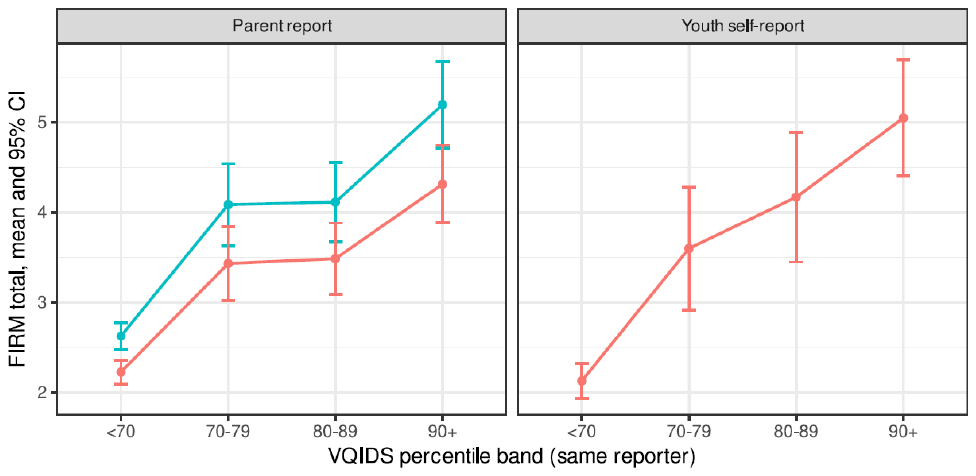

**Tables S1–S4. Measurement**-**invariance (DIF) details**

Each table reports both detection methods (the likelihood-ratio test statistics and the ordinal-logistic-regression effect size) and the adjudicated decision. In every contrast, no item met the practical threshold, so none is retained as differential item functioning.

**Table S1.**

Sex DIF based on Parent report

|  | *X^2^* | *df* | *p* | adj_*p* | AIC | BIC | SIDS | UIDS | ESSD | *lordif* Δ*R²* | DIF (adj.) |
| --- | --- | --- | --- | --- | --- | --- | --- | --- | --- | --- | --- |
| Sad Mood | 8.580 | 4.000 | 0.072 | 0.290 | −0.580 | 22.294 | 0.000 | 0.000 | 0.000 | 0.001 | - |
| Impaired concentration/  decision making | 14.085 | 4.000 | 0.007 | 0.035 | −6.085 | 16.789 | 0.087 | 0.087 | 0.252 | 0.003 | - |
| Negative self-outlook | 3.965 | 4.000 | 0.411 | 0.630 | 4.035 | 26.910 | 0.000 | 0.000 | 0.000 | 0.000 | - |
| Reduced involvement | 4.741 | 4.000 | 0.315 | 0.630 | 3.259 | 26.134 | 0.000 | 0.000 | 0.000 | 0.001 | - |
| Low energy | 6.066 | 4.000 | 0.194 | 0.583 | 1.934 | 24.809 | 0.000 | 0.000 | 0.000 | 0.001 | - |

*Note:* X^2^/*df*/*p* = likelihood-ratio (*mirt*) DIF test; adj_*p* = Benjamini-Hochberg adjusted. SIDS/UIDS = signed/unsigned expected-score difference (raw points); ESSD = expected-score standardized difference. *lordif* Δ*R²* = McFadden pseudo-*R²* change (the second, ordinal-logistic-regression method; flagged at Δ*R²* ≥ 0.02); DIF (adj.) = adjudicated decision, retained only if the raw-point effect size reaches 0.2.

**Table S2.**

Age DIF based on Parent report: developmental band (6-11 vs 12-18)

|  | *X^2^* | *df* | *p* | adj_*p* | AIC | BIC | SIDS | UIDS | ESSD | *lordif* Δ*R²* | DIF (adj.) |
| --- | --- | --- | --- | --- | --- | --- | --- | --- | --- | --- | --- |
| Sad Mood | 1.615 | 4.000 | 0.806 | 1.000 | 6.385 | 29.259 | 0.000 | 0.000 | 0.000 | 0.000 | - |
| Impaired concentration/  decision making | 33.848 | 4.000 | 0.000 | 0.000 | −25.848 | −2.973 | 0.124 | 0.124 | 0.379 | 0.009 | - |
| Negative self-outlook | 6.898 | 4.000 | 0.141 | 0.424 | 1.102 | 23.977 | 0.000 | 0.000 | 0.000 | 0.001 | - |
| Reduced involvement | 1.692 | 4.000 | 0.792 | 1.000 | 6.308 | 29.183 | 0.000 | 0.000 | 0.000 | 0.000 | - |
| Low energy | 39.450 | 4.000 | 0.000 | 0.000 | −31.450 | −8.576 | −0.079 | 0.079 | −0.284 | 0.011 | - |

*Note:* X^2^/*df*/*p* = likelihood-ratio (*mirt*) DIF test; adj_*p* = Benjamini-Hochberg adjusted. SIDS/UIDS = signed/unsigned expected-score difference (raw points); ESSD = expected-score standardized difference. *lordif* Δ*R²* = McFadden pseudo-*R²* change (the second, ordinal-logistic-regression method; flagged at Δ*R²* ≥ 0.02); DIF (adj.) = adjudicated decision, retained only if the raw-point effect size reaches 0.2.

**Table S3.**

Youth self-report: sex

|  | *X^2^* | *df* | *p* | adj_*p* | AIC | BIC | SIDS | UIDS | ESSD | *lordif* Δ*R²* | DIF (adj.) |
| --- | --- | --- | --- | --- | --- | --- | --- | --- | --- | --- | --- |
| Sad Mood | 1.985 | 4.000 | 0.739 | 0.742 | 6.015 | 25.670 | 0.000 | 0.000 | 0.000 | 0.001 | - |
| Impaired concentration/  decision making | 5.414 | 4.000 | 0.247 | 0.742 | 2.586 | 22.241 | 0.000 | 0.000 | 0.000 | 0.001 | - |
| Negative self-outlook | 10.109 | 4.000 | 0.039 | 0.193 | −2.109 | 17.546 | 0.000 | 0.000 | 0.000 | 0.003 | - |
| Reduced involvement | 6.626 | 4.000 | 0.157 | 0.628 | 1.374 | 21.029 | 0.000 | 0.000 | 0.000 | 0.001 | - |
| Low energy | 4.596 | 4.000 | 0.331 | 0.742 | 3.404 | 23.059 | 0.000 | 0.000 | 0.000 | 0.001 | - |

*Note:* X^2^/*df*/*p* = likelihood-ratio (*mirt*) DIF test; adj_*p* = Benjamini-Hochberg adjusted. SIDS/UIDS = signed/unsigned expected-score difference (raw points); ESSD = expected-score standardized difference. *lordif* Δ*R²* = McFadden pseudo-*R²* change (the second, ordinal-logistic-regression method; flagged at Δ*R²* ≥ 0.02); DIF (adj.) = adjudicated decision, retained only if the raw-point effect size reaches 0.2.

**Table S4.**

Youth self-report: within-adolescence band (12-14 vs 15-18)

|  | *X^2^* | *df* | *p* | adj_*p* | AIC | BIC | SIDS | UIDS | ESSD | *lordif* Δ*R²* | DIF (adj.) |
| --- | --- | --- | --- | --- | --- | --- | --- | --- | --- | --- | --- |
| Sad Mood | 3.331 | 4.000 | 0.504 | 1.000 | 4.669 | 24.324 | 0.000 | 0.000 | 0.000 | 0.001 | - |
| Impaired concentration/  decision making | 12.423 | 4.000 | 0.014 | 0.072 | −4.423 | 15.232 | 0.000 | 0.000 | 0.000 | 0.006 | - |
| Negative self-outlook | 9.637 | 4.000 | 0.047 | 0.188 | −1.637 | 18.018 | 0.000 | 0.000 | 0.000 | 0.000 | - |
| Reduced involvement | 2.585 | 4.000 | 0.629 | 1.000 | 5.415 | 25.070 | 0.000 | 0.000 | 0.000 | 0.001 | - |
| Low energy | 6.066 | 4.000 | 0.194 | 0.583 | 1.934 | 21.589 | 0.000 | 0.000 | 0.000 | 0.002 | - |

*Note:* X^2^/*df*/*p* = likelihood-ratio (*mirt*) DIF test; adj_*p* = Benjamini-Hochberg adjusted. SIDS/UIDS = signed/unsigned expected-score difference (raw points); ESSD = expected-score standardized difference. *lordif* Δ*R²* = McFadden pseudo-*R²* change (the second, ordinal-logistic-regression method; flagged at Δ*R²* ≥ 0.02); DIF (adj.) = adjudicated decision, retained only if the raw-point effect size reaches 0.2.

**Table S5. Robustness to the sparse top category**

Refitting the item model after merging the seldom-used top response category left the item properties and each child’s estimated level essentially unchanged, confirming the results are not an artifact of that rare category.

**Table S5.**

Top-category robustness (categories 2 and 3 collapsed)

|  | a (full) | a (collapsed) | Δ |
| --- | --- | --- | --- |
| Parent report | | | |
| Sad Mood | 2.04 | 2.05 | 0.01 |
| Impaired concentration/decision making | 1.50 | 1.48 | −0.03 |
| Negative self-outlook | 2.33 | 2.44 | 0.11 |
| Reduced involvement | 2.19 | 2.15 | −0.04 |
| Low energy | 2.00 | 1.99 | −0.02 |
| Youth self-report | | | |
| Sad Mood | 2.57 | 2.51 | −0.06 |
| Impaired concentration/decision making | 1.94 | 1.93 | −0.01 |
| Negative self-outlook | 2.17 | 2.33 | 0.15 |
| Reduced involvement | 2.03 | 2.03 | 0.00 |
| Low energy | 2.06 | 1.95 | −0.10 |

*Note:* a_full / a_collapsed = item discrimination before / after collapsing the sparse top category; delta = collapsed - full. Latent-score (theta) rank stability: Parent report r = 0.999, rho = 1.000; Youth self-report r = 0.998, rho = 0.999.

**Table S6. Raw**-**score versus model**-**based scoring of the norms**

As a precision check, we also derived the norms from a model-based (item response theory) score instead of the raw total. The two approaches classified children near-identically (agreement of about 98% at the 90th-percentile flag), so the simpler raw-score norms are used throughout.

**Table S6.**

Agreement between raw-score and model-based model

|  | T-score agreement | | % classified alike at cutoff | | |
| --- | --- | --- | --- | --- | --- |
|  | T *r* | T ρ | P70 | P80 | P90 |
| Parent report | 0.978 | 0.983 | 91.5 | 94.1 | 98.1 |
| Youth self-report | 0.983 | 0.989 | 91.3 | 94.4 | 98.4 |

*Note:* T = normalized T-score (50 + 10 z) from each norming model. r = Pearson (linear) agreement; ρ = Spearman (rank) agreement between the raw-score and model-based T-scores. The cutoff columns give the % of cases classified identically (above vs below) by the two norming approaches at each percentile flag.

**S7. How the norming model was chosen**

The VQIDS total is a count: a whole number from 0 to 15, with many zeros and a long right tail. A normal-curve model is therefore inappropriate. We restricted the candidates in advance to three distributions built for exactly this kind of variable, each fitted with the identical predictor structure (sex, a smooth function of age, and a sex-by-age smooth, with the spread also free to change with age):

- **NBI**, the negative binomial, which lets the spread exceed what a simple count model allows (over-dispersion);
- **ZINBI**, the same model plus a separate extra probability of scoring zero (zero-inflation);
- **ZANBI**, a hurdle model, which treats “any symptoms at all” and “how many” as two distinct processes.

The three were compared by BIC, which rewards fit and penalizes complexity (Table S7). **For both reporters the plain NBI won.** This is the substantively interesting result: the negative binomial already absorbs the large number of zeros on its own, so adding a dedicated zero-inflation or hurdle component costs extra parameters without improving fit. The floor in the VQIDS is not a separate phenomenon needing its own machinery; it falls out naturally from an over-dispersed count.

**Table S7.**

Comparison of candidate count distribution for the norms

|  | df | BIC | Delta BIC |
| --- | --- | --- | --- |
| Parent report | | | |
| NBI (negative binomial) | 5.0 | 8,321.9 | 0.0 |
| ZINBI (zero-inflated) | 7.0 | 8,334.7 | 12.8 |
| ZANBI (hurdle) | 6.3 | 8,353.1 | 31.1 |
| Youth self-report | | | |
| NBI (negative binomial) | 5.4 | 4,032.7 | 0.0 |
| ZINBI (zero-inflated) | 7.6 | 4,044.7 | 12.0 |
| ZANBI (hurdle) | 11.0 | 4,111.4 | 78.7 |

*Note:* All three models share the same predictors (sex, smooth age, sex-by-age smooth; spread free to vary with age) and differ only in the distribution. BIC rewards fit and penalizes complexity; lower is better. Delta BIC is the difference from the best model within each reporter. NBI is retained for both.

Diagnostics for the retained model confirm that it fits across the whole age range. The worm plots (Figures S2 and S3) show the residuals within four age windows: points lying close to the horizontal line and inside the confidence bands indicate the model has captured the location, spread, and shape of the score distribution at each age. Tables S8 and S9 report the correlations among the fitted model’s parameters. The check that matters here is whether the part of the model governing the typical score and the part governing the spread are estimated independently of one another, and they are: those cross-correlations are all close to zero. (The strong negative correlations between an intercept and its own age term are an expected consequence of the parameterization and carry no diagnostic weight.).

**Figure S2**

*Worm plot for the retained parent-report model (NBI), across four age windows. Points within the bands indicate adequate fit at each age.*

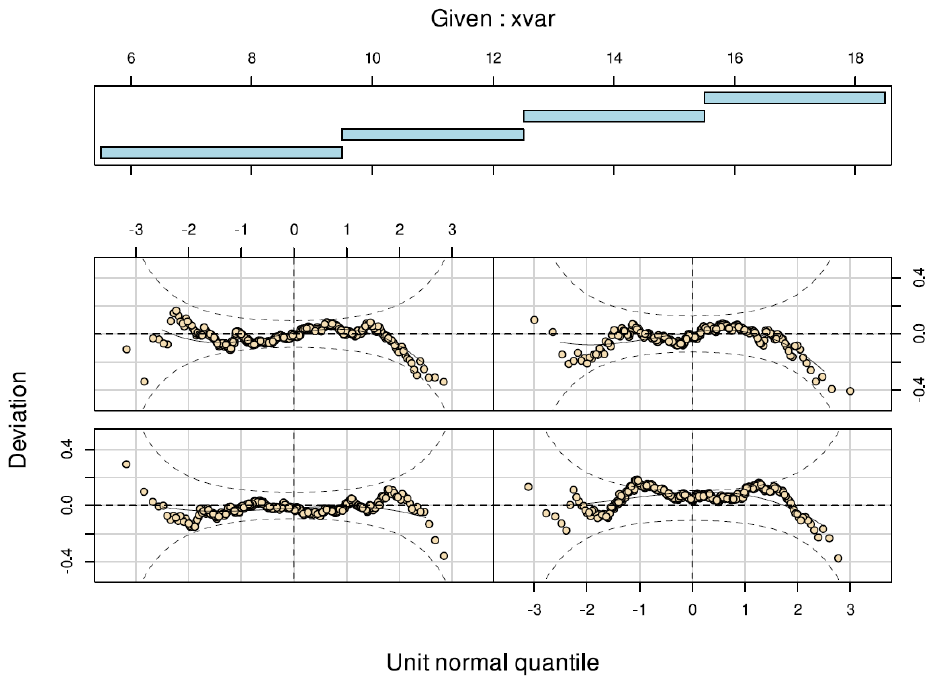

**Figure S3**

*Worm plot for the retained youth self-report model (NBI), across four age windows.*

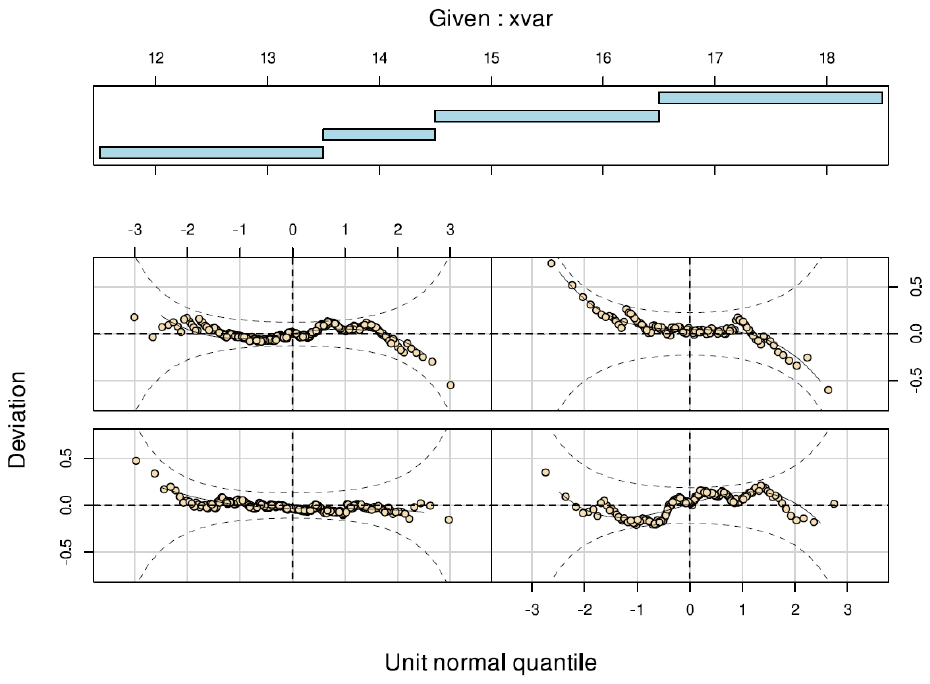

**Table S8.**

Parent report: parameter correlations (NBI)

|  | (Intercept) | femaleyn1 | pb(actual_age) | (Intercept).1 | pb(actual_age).1 |
| --- | --- | --- | --- | --- | --- |
| (Intercept) | 1.00 | −0.17 | −0.93 | 0.01 | −0.01 |
| femaleyn1 | −0.17 | 1.00 | −0.10 | 0.12 | −0.13 |
| pb(actual_age) | −0.93 | −0.10 | 1.00 | −0.04 | 0.04 |
| (Intercept).1 | 0.01 | 0.12 | −0.04 | 1.00 | −0.97 |
| pb(actual_age).1 | −0.01 | −0.13 | 0.04 | −0.97 | 1.00 |

*Note:* Correlations among the estimated parameters of the retained NBI model (mu = the typical score; sigma = the spread). The large negative values *within* a block (intercept versus the age term) are an expected consequence of the parameterization, not a problem. The informative entries are the *cross-block* correlations between the mu and sigma terms, which are all near zero: the location and spread components of the model are estimated independently rather than trading off against one another.

**Table S9.**

Youth self-report: parameter correlations (NBI)

|  | (Intercept) | femaleyn1 | pb(actual_age) | (Intercept).1 | pb(actual_age).1 |
| --- | --- | --- | --- | --- | --- |
| (Intercept) | 1.00 | −0.05 | −0.99 | 0.03 | −0.03 |
| femaleyn_y1 | −0.05 | 1.00 | −0.07 | 0.02 | −0.02 |
| pb(actual_age) | −0.99 | −0.07 | 1.00 | −0.03 | 0.03 |
| (Intercept).1 | 0.03 | 0.02 | −0.03 | 1.00 | −0.99 |
| pb(actual_age).1 | −0.03 | −0.02 | 0.03 | −0.99 | 1.00 |

*Note:* Correlations among the estimated parameters of the retained NBI model (mu = the typical score; sigma = the spread). The large negative values *within* a block (intercept versus the age term) are an expected consequence of the parameterization, not a problem. The informative entries are the *cross-block* correlations between the mu and sigma terms, which are all near zero: the location and spread components of the model are estimated independently rather than trading off against one another.

**Table S10. Parent–youth agreement on individual FIRM items**

Item-level agreement for the 20 grid items. Because most items describe rare facts, kappa should be read alongside prevalence: when a fact is rare, kappa can be low even though the two informants give the same answer for nearly every family (the base-rate paradox).

**Table S10a.**

Item-level parent-youth agreement on the FIRM grid

|  | Parent % yes | Youth % yes | % agreement | Kappa |
| --- | --- | --- | --- | --- |
| Suicide attempt / Grandparents | 6.4 | 4.7 | 95.0 | 0.53 |
| Suicide attempt / Parents | 8.4 | 9.3 | 92.0 | 0.50 |
| Suicide attempt / Aunts/uncles | 8.2 | 7.1 | 93.2 | 0.52 |
| Suicide attempt / Siblings | 3.4 | 4.4 | 96.2 | 0.49 |
| Substance problems / Grandparents | 27.3 | 22.1 | 83.5 | 0.56 |
| Substance problems / Parents | 26.7 | 24.8 | 86.0 | 0.63 |
| Substance problems / Aunts/uncles | 26.7 | 24.3 | 81.3 | 0.51 |
| Substance problems / Siblings | 5.1 | 5.8 | 93.8 | 0.39 |
| Psychiatric hospitalization / Grandparents | 7.8 | 7.4 | 93.8 | 0.56 |
| Psychiatric hospitalization / Parents | 11.5 | 12.0 | 91.8 | 0.60 |
| Psychiatric hospitalization / Aunts/uncles | 9.0 | 8.8 | 91.1 | 0.45 |
| Psychiatric hospitalization / Siblings | 5.4 | 5.3 | 96.5 | 0.65 |
| Depression / Grandparents | 28.6 | 24.2 | 83.3 | 0.57 |

*Note:* n = 1008 dyads. Cohen's kappa on the 1 = endorsed / 0 = not coding. Items are labeled problem area / relative class.

**Table S10b.**

Item-level parent-youth agreement on the FIRM grid

|  | Parent % yes | Youth % yes | % agreement | Kappa |
| --- | --- | --- | --- | --- |
| Depression / Parents | 43.4 | 41.0 | 83.5 | 0.66 |
| Depression / Aunts/uncles | 26.0 | 22.8 | 80.4 | 0.47 |
| Depression / Siblings | 13.8 | 13.1 | 90.4 | 0.59 |
| Mania/bipolar / Grandparents | 10.8 | 8.9 | 91.6 | 0.53 |
| Mania/bipolar / Parents | 14.7 | 15.0 | 92.2 | 0.69 |
| Mania/bipolar / Aunts/uncles | 11.8 | 10.0 | 90.7 | 0.52 |
| Mania/bipolar / Siblings | 4.5 | 4.5 | 95.4 | 0.47 |

*Note: N* = 1008 dyads. Cohen's kappa on the 1 = endorsed / 0 = not coding. Items are labeled problem area / relative class.

**Table S11. FIRM discrimination at graded VQIDS benchmarks**

AUROCs for both FIRM totals at the 70th, 80th, and 90th percentile benchmarks, each reporter predicting its own benchmark on the widest classifiable sample. The parent rows therefore use the full parent sample, not the dyad subset, so they differ slightly from the matched-dyad matrix in Table 10, which is the comparison to rely on when weighing reporters against each other.

**Table S11.**

FIRM discrimination of graded VQIDS benchmark (same-reporter, widest samples)

| Benchmark | Total | AUROC | n flagged | n |
| --- | --- | --- | --- | --- |
| Parent report | | | | |
| ≥ 90th percentile | Grid | .667 [.631, .704] | 236 | 2252 |
| ≥ 90th percentile | Full | .686 [.650, .721] | 236 | 2252 |
| ≥ 80th percentile | Grid | .649 [.621, .676] | 475 | 2252 |
| ≥ 80th percentile | Full | .660 [.633, .687] | 475 | 2252 |
| ≥ 70th percentile | Grid | .653 [.629, .678] | 674 | 2252 |
| ≥ 70th percentile | Full | .665 [.641, .689] | 674 | 2252 |
| Youth self-report | | | | |
| ≥ 90th percentile | Grid | .731 [.682, .780] | 106 | 1008 |
| ≥ 80th percentile | Grid | .712 [.671, .752] | 195 | 1008 |
| ≥ 70th percentile | Grid | .697 [.661, .733] | 290 | 1008 |

*Note:* AUROC [95% CI]; 0.5 = chance. Same-reporter analyses: these include shared-rater variance and are upper bounds (see Table 13 for the cross-informant tests). Grid and full totals are shown for parent report; youth self-report is the grid total only, because the youth form's two extra items ask about the adolescent's own diagnoses rather than a relative's.

**Table S12.**

VQIDS and FIRM ROC/AUC Summary, Classifying Parent-Reported and Youth-Reported Depression Diagnosis (N = 1008, *except parent full sample paired with parent reported diagnosis, *N*=2251)

| Statistic | Parent Reported Dx | Youth Reported Dx (*N* = 1008) |
| --- | --- | --- |
| **VQIDS** | | |
| Parent (full sample) | AUC* = .766 [.730, .801] | AUC = .766 [.729, .803] |
| Parent (dyads) | AUC = .738 [.693, .782] | AUC = .766 [.729, .803] |
| Youth | AUC = .756 [.712, .801] | AUC = .775 [.738, .813] |
| **FIRM** | | |
| Parent (full sample) | AUC* = .760 [.727, .794] | AUC = .715 [.676, .755] |
| Parent (dyads) | AUC = .736 [.694, .778] | AUC = .715 [.676, .755] |
| Youth | AUC = .768 [.729, .807] | AUC = .775 [.739, .811] |

**Table S13.**

FIRM ROC/AUC Summary, Classifying Parent-Reported and Youth-Reported Bipolar Diagnosis (*N* = 1008 except for parent FIRM paired with parent reported diagnosis, *N* = 2252)

| Statistic | Parent Reported Dx | Youth Reported Dx |
| --- | --- | --- |
| Parent (full sample) | AUC* = .743 [.652, .834] | AUC = .797 [.732, .862] |
| Parent (dyads) | AUC = .727 [.599, .855] | AUC = .797 [.732, .862] |
| Youth | AUC = .794 [.708, .880] | AUC = .831 [.781, .881] |

**Table S14.**

Parent-Youth Categorical Agreement on Diagnosis Status: Depression and Bipolar Disorder (*N* = 1,008 matched dyads)

| Statistic | Depression | Bipolar |
| --- | --- | --- |
| Parent ‘Yes’ / Youth ‘Yes’ (both agree) | 111 | 11 |
| Parent ‘No’ / Youth ‘Yes’ (youth only) | 66 | 24 |
| Parent ‘Yes’ / Youth ‘No’ (parent only) | 28 | 4 |
| Parent ‘No’ / Youth ‘No’ (both agree) | 803 | 969 |
| Observed (raw) agreement | .907 | .972 |
| Cohen’s kappa [95% CI] | .65 [.58, .71] | .43 [.26, .60] |
| Gwet’s AC1 [95% CI] | .873 [.847, .900] | .971 [.960, .982] |
| Prevalence: parent / youth | .138 / .176 | .015 / .035 |

**Table S15.**

Model Performance: VQIDS and FIRM (Parent and Youth) Predicting Four Diagnostic Outcomes (*N*=1,008)

| **Model** | **AUC [95% CI]** | | **Brier** | **Spiegelhalter *Z* (*p*)** |
| --- | --- | --- | --- | --- |
| **Depression, parent-reported** | | | | |
| VQIDS (own = parent) | .738 [.693,.782] | | .1065 | 0.183 (.855) |
| FIRM (own = parent) | .736 [.694,.778] | | .1090 | 0.225 (.822) |
| VQIDS (other = youth) | .756 [.712,.801] | | .1036 | 0.143 (.887) |
| FIRM (other = youth) | .768 [.729,.807] | | .1066 | 0.357 (.721) |
| Within-informant combined | .792 [.754,.830] | | .1003 | 0.266 (.791) |
| Cross-informant combined | .808 [.771,.845] | | .0975 | 0.260 (.795) |
| All four scales combined | .819 [.784,.854] | | .0951 | 0.253 (.800) |
| **Depression, youth-reported** | | | | |
| VQIDS (own = youth) | .775 [.738,.813] | | .1199 | 0.204 (.838) |
| FIRM (own = youth) | .775 [.739,.811] | | .1265 | 0.464 (.643) |
| VQIDS (other = parent) | .766 [.729,.803] | | .1258 | 0.385 (.700) |
| FIRM (other = parent) | .715 [.676,.755] | | .1357 | 0.301 (.763) |
| Within-informant combined | .828 [.797,.860] | | .1118 | 0.391 (.696) |
| Cross-informant combined | .800 [.767,.832] | | .1213 | 0.491 (.624) |
| All four scales combined | .840 [.811,.869] | | .1094 | 0.425 (.671) |
| **Bipolar, parent-reported** | | | | |
| VQIDS (own = parent) | .799 [.707,.891] | | .0144 | 0.056 (.956) |
| FIRM (own = parent) | .727 [.599,.855] | | .0145 | 0.023 (.982) |
| VQIDS (other = youth) | .718 [.588,.848] | | .0144 | 0.000 (1.000) |
| FIRM (other = youth) | .794 [.708,.880] | | .0144 | 0.033 (.974) |
| Within-informant combined | .821 [.717,.925] | | .0142 | 0.065 (.948) |
| Cross-informant combined | .798 [.705,.890] | | .0142 | 0.010 (.992) |
| All four scales combined | .832 [.740,.924] | | .0140 | 0.022 (.983) |
| **Bipolar, youth-reported** | | | | |
| VQIDS (own = youth) | .817 [.754,.881] | | .0317 | 0.180 (.857) |
| FIRM (own = youth) | .831 [.781,.881] | | .0324 | 0.261 (.794) |
| VQIDS (other = parent) | .808 [.751,.865] | | .0327 | 0.241 (.810) |
| FIRM (other = parent) | .797 [.732,.862] | | .0327 | 0.209 (.835) |
| Within-informant combined | .863 [.814,.913] | | .0307 | 0.278 (.781) |
| Cross-informant combined | .842 [.781,.903] | | .0315 | 0.288 (.773) |
| All four scales combined |  | .871  [.821, .921] | .0301 | 0.240 (.810) |

*Note*: Both family history and brief screening tools are recommended first line assessment strategies. Logistic regressions and calibration statistics (Brier, and Spiegelhalter’s *Z*) assessed how the VQIDS and FIRM did predicting the youth diagnoses of mood disorders, both within informant and across informant.
